# ADVANCED ARTIFICIAL INTELLIGENCE ENABLED METHODS FOR EARLY DETECTION OF NON-ALCOHOLIC FATTY LIVER DISEASE AND ASSOCIATED HEALTH RISKS

**DOI:** 10.64898/2026.02.12.26345169

**Authors:** N Sastha Kumar, Gokul Krishna Sreekumar, Hari Krishnan, Sai Janakiram Chinnakanu, M Nidheesh, Sabarinath Subramaniam

## Abstract

Non-alcoholic fatty liver disease (NAFLD) is a globally prevalent hepatic condition caused by the buildup of fat in the liver. It is frequently associated with metabolic comorbidities such as hypertension, cardiovascular disease (CVD), and prediabetes. However, early detection remains challenging due to the asymptomatic progression, and existing primary diagnostic methods, such as imaging or liver biopsy, are often expensive and inaccessible in rural areas. This study proposes a two-stage, interpretable machine learning pipeline for the non-invasive and cost-effective prediction of NAFLD and its key comorbidities using routine clinical parameters.

The NAFLD prediction model was developed using the XGBoost algorithm, trained on a hybrid dataset that combines real patient data with rule-based synthetic data generated by simulating clinically plausible cases. Upon NAFLD-positive prediction, three separate XGB models, trained on data labelled based on thresholds, assess individual risks for hypertension, cardiovascular disease, and prediabetes. Explainability is obtained using SHAP (SHapley Additive exPlanations), which provides insight into feature relevance, while biomarker radar plots help in the visual interpretation of comorbidities. A user-friendly Streamlit interface enables real-time interaction with the tool for potential clinical application.

The NAFLD model demonstrated robust performance, while the models used for predicting comorbidities achieved perfect performance, which may be a reflection of the limited dataset size used in the second stage. This work underscores the potential of AI-driven tools in NAFLD diagnosis, particularly when combined with explainable AI methods.

## 1. INTRODUCTION

Non-alcoholic fatty liver disease is one of the most common chronic liver conditions worldwide, affecting more than one-third of the total population. It is caused by the accumulation of excessive hepatic fat in patients with little to no alcoholism. Regional variations show that the Middle East and South America are at very high risk of the disease[1,2]. NAFLD ranges from simple steatosis (a fat buildup in the hepatocytes) to more serious stages like non-alcoholic steatohepatitis (NASH), fibrosis, cirrhosis, and hepatocellular carcinoma (HCC). It is reversible in early stages through a combination of medication and lifestyle changes, but the major threat of the disease lies in its potential to progress undetected; by the time the patient decides to get a check-up, it has often progressed to severe, irreversible stages requiring liver transplantation [3]. The disease comes with massive financial burdens, and the economic impact of NAFLD in the US alone is $103 billion per year [4].

NAFLD is not an isolated hepatic condition but a multi-system disease linked to multiple comorbidities, which compounds its clinical impact beyond the liver. Comorbidities associated with NAFLD include cardiovascular disease (CVD), hypertension, prediabetes, etc., which accelerate the progression of the disease and thereby increase the risk of mortality in patients. CVD is a broad term for diseases affecting the heart and blood vessels, and it remains the leading cause of mortality in NAFLD patients [5]. Prediabetes precedes Type 2 Diabetes and is seen during the early stages of NAFLD [6]. Co-prevalence of Diabetes reaches up to 60% and it poses a higher risk of advanced fibrosis and hepatocellular carcinoma [7]. Hypertension is a condition in which the blood pressure remains consistently high. It is seen in 40% of NAFLD patients and increases the risk of cardiovascular mortality [8].

Current diagnostic strategies for NAFLD include liver biopsy, considered the gold standard, as well as imaging methods such as ultrasonography and magnetic resonance imaging. Biopsy is invasive and risks minor injuries while being expensive at over 1000 USD per procedure. It is impractical for population-level screening. Imaging methods are also comparatively costly and require specialized equipment, which might limit their availability in rural areas. The presence of obesity has also been observed to decrease ultrasound image clarity [9,10]. This has created a need for the use of non-invasive diagnostic methods, particularly within primary care, and for clinical data that includes blood biomarkers, demographics, anthropometrics, and blood pressure, which can be collected easily through routine tests that are considerably less expensive than current methods [11–16].

Various studies have demonstrated that Artificial Intelligence (AI) models are capable of extracting complex patterns and relationships from vast amounts of data [9,10]. Machine Learning (ML) algorithms trained on routine clinical data for diagnosing NAFLD offer the potential to replace invasive techniques. They can be used as a clinical aid to flag at-risk patients in a faster, cheaper way that is also accessible in all regions. It also offers the advantage of combining both raw and derived features to strengthen the prediction. Derived features, such as the Triglyceride-Glucose index (TyG), Metabolic score for Insulin Resistance (METS-IR), and Triglyceride to High-density lipoprotein ratio (TG/HDL), have been observed to be reliable indicators of NAFLD [17–21], and they can be easily calculated using standard clinical parameters. In addition to diagnosing NAFLD, it is also essential to address the comorbidities that often co-occur with it. Since NAFLD shares bidirectional relationships with conditions such as CVD, Prediabetes, and Hypertension, it is possible to create a diagnostic pipeline that checks for NAFLD and each of these comorbidities in patients. Clinically, the comorbidities are measured through specific feature thresholds, obtained through clinical tests. The ability of ML models to learn from large, diverse datasets [22] makes them suitable for multi-morbidity prediction.

The primary concern in adopting AI tools is their ‘black-box’ nature, which limits their interpretability and may lead clinicians to approach the outputs with skepticism. Therefore, explainability is a crucial concept in AI- and ML-driven diagnosis. Explainable AI techniques, such as SHAP (Shapley Additive Explanations), provide feature-level attributions that clarify why a model made a specific prediction [23]. SHAP works on the principle of game theory by assigning credit to each feature that contributes to a model’s prediction to make it more interpretable. It provides graphical representations of how much each feature contributes to the prediction, which can increase confidence in adopting the models by addressing transparency issues. Additionally, methods like biomarker radar plots can be used, with the threshold features marked on the radar, to identify which features show abnormality.

This study aims to develop an interpretable and user-friendly diagnostic tool that can serve as a primary diagnostic aid to predict the probability of a patient developing NAFLD and its related comorbidities, utilizing readily available clinical data as a non-invasive, cost-effective, and widely accessible alternative to existing diagnostic methods. The approach employs a two-stage pipeline; the first stage predicts NAFLD, and the second stage identifies comorbidities in patients who test positive for NAFLD. XGB models were selected for prediction, and SHAP and radar plots were used to address explainability. The pipeline was integrated into a web-based application built using Streamlit. This Python library enables users to build and share web applications, providing an intuitive interface for users to input patient data and receive predictions along with corresponding visual explanations.

## 2. MATERIALS AND METHODS

### 2.1 Study Design

This study follows a two-stage pipeline aimed at detecting Non Alcoholic Fatty Liver Disease and three of its prevalent comorbidities using ML models trained on clinical data and additional synthetic data generated through rule-based methods to ensure class balance. Hypertension, Prediabetes, and Cardiovascular disease were selected as comorbidities as they were observed to be commonly occurring in NAFLD patients and had the potential to be detected based on routinely collected, non-invasive, and inexpensive clinical data, unlike other comorbidities like sleep apnea or chronic kidney disease (CKD), which needed non-routine test data. The design involves data collection, preprocessing, synthetic data augmentation, model development, evaluation, explainability, and deployment into a user-friendly interface.

### 2.2 Data Collection

#### 2.2.1 Dataset for NAFLD Prediction

The primary dataset was obtained from a previous study that focused on advancing the predictive comparison of non-alcoholic fatty liver disease [24]. The data were taken from the Dryad database, a repository funded by the National Science Foundation. They were collected as part of a study on ectopic fat, obesity, and type 2 diabetes mellitus [25]. The dataset comprised 14,913 records and 23 features, with NAFLD status categorized as either negative or positive based on abdominal ultrasound results. The dataset included demographic features like sex and age, anthropometric data like body mass index (BMI), waist circumference (WC), visceral fat status and obesity status, blood biomarkers like alanine aminotransferase (ALT), aspartate aminotransferase (AST), gamma-glutamyl transferase (GGT), high-density lipoprotein (HDL), total cholesterol (TC), triglycerides (TG), fasting plasma glucose (FPG), hemoglobin A1c (HbA1c), blood pressure data like systolic blood pressure (SBP) and diastolic blood pressure (DBP) and derived features like TyG Index, TyG-BMI, TyG-WC, TG/HDL ratio and METS-IR. Derived features were retained because the study from which the dataset was obtained identified them as key predictors. No missing values were observed in the dataset.

#### 2.2.2 Dataset for Predicting Comorbidities

A subset of NAFLD-positive entries, which comprised a total of 2,620 records, was extracted from the Dryad dataset for predicting prediabetes, CVD, and hypertension that occurred in NAFLD patients. Binary labels were calculated for each comorbidity based on their clinical thresholds, given as follows for positive cases [26,27]; Hypertension: SBP ≥ 130 mmHg or DBP ≥ 80mmHg, Prediabetes: FPG 5.6-6.9 mmol/L (100–125 mg/dL) or HbA1c 5.7–6.4% and CVD: TyG ≥ 8.5 or TG/HDL ≥ 3.5 or SBP ≥ 130 mmHg or DBP ≥ 80mmHg.

Table 1 summarizes the distribution of each comorbidity after labelling.

**TABLE 1:** Distribution of comorbidities in the dataset.

| Comorbidity | Positive Cases | Negative Cases |
| --- | --- | --- |
| Hypertension | 1140 | 1480 |
| CVD | 1793 | 827 |
| Prediabetes | 1047 | 1573 |

### 2.3 Data Preprocessing

The derived features included in the dataset were observed to contain inconsistencies in unit conversion. Raw biomarkers were reported in millimoles per liter (mmol/L), but derived features were calculated in milligrams per deciliter (mg/dL) units. To address this, all derived features were recalculated with consistent units (mmol/L) and correct clinical formulas. For the TyG Index, which involves TG and FPG, the original equation uses milligrams per deciliter (mg/dL) units. Since we are considering mmol/L (the commonly used unit outside the US), we needed to convert the mmol/L units of the raw features to match the mg/dL unit expected in the equation. For this purpose, TG values in mmol/L are multiplied by 88.57, and FPG values in mmol/L are multiplied by 18, which adds a constant of 1594.26 (88.57 × 18) to the equation used to calculate TyG Index, when raw features are in mmol/L. Similarly, METS-IR also involves TG, FPG, and HDL (where values in mmol/L are multiplied by 38.67 to convert to mg/dL); for calculating this, an internal conversion was applied, where each value is converted from mmol/L to mg/dL before being substituted into the equation. The mathematical equations of all indices used in this study are provided in Figure 1.

**Figure 1.**
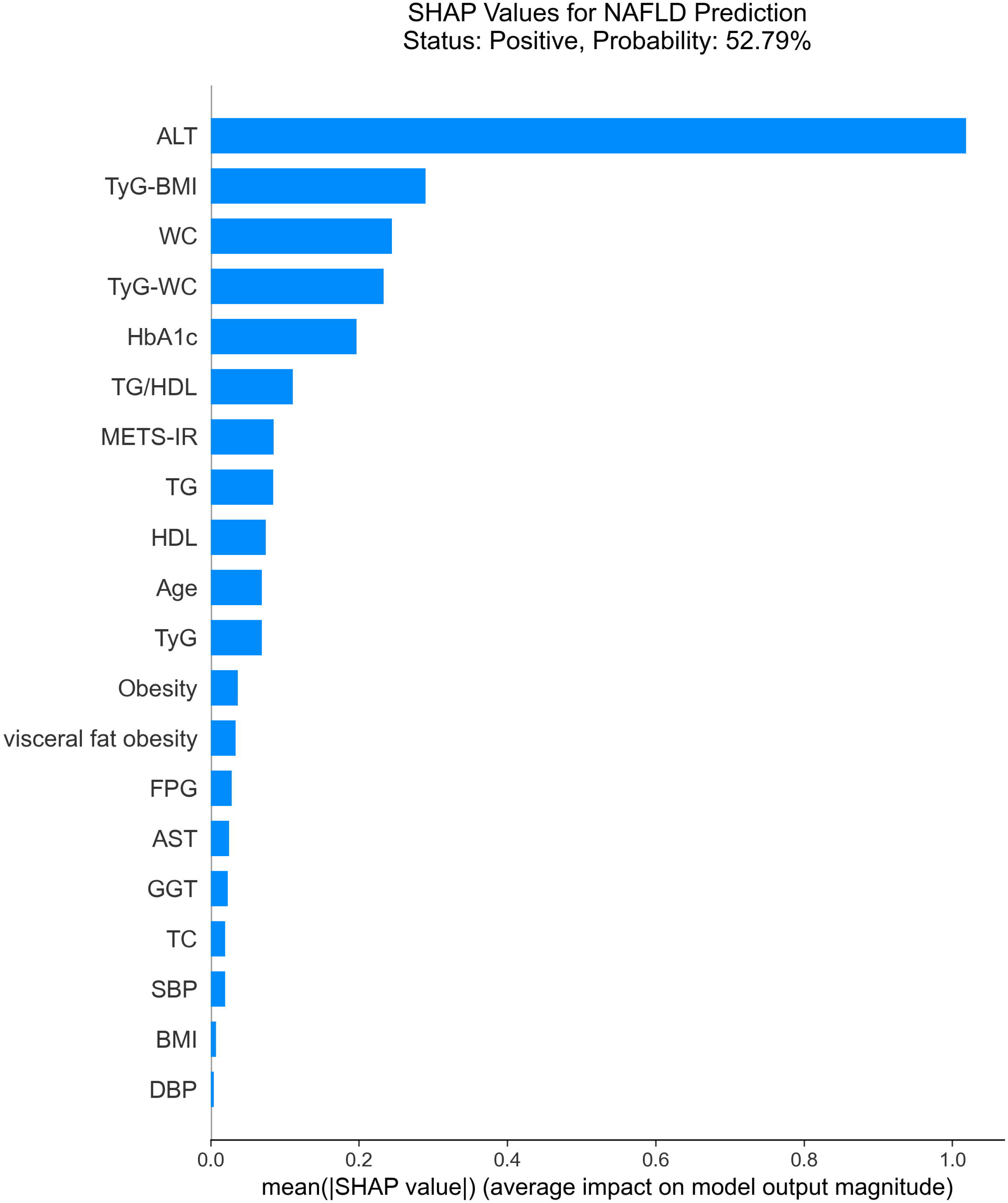
Mathematical equations of metabolic indices used in the study. All indices were computed using standardized formulas following internal conversion from mmol/L to mg/dL where applicable.

StandardScaler, a function that standardizes features by removing the mean and scaling to unit variance, was used for feature scaling before training the model to ensure uniform contribution of each variable in the model.

### 2.4 Synthetic Data Generation

The dataset for NAFLD prediction had significant class imbalance (2620 positive cases vs 12,293 negative cases). To address this, synthetic data (10,000 samples) was generated using a rule-based method where data was synthesized by sampling random values within the clinical range of each feature, as observed in the actual dataset. This ensured that the generated values stayed within a realistic clinical range. Derived features for these samples were calculated post-synthesis using the appropriate formulas (Section 2.3). Synthetic data was not generated for the comorbidity dataset due to its limited size and associated higher risk of generating samples that deviate from real-world patterns.

### 2.5 Model Development and Evaluation

#### 2.5.1 Stage 1: NAFLD Prediction

Three algorithms - logistic regression (LR), random forest (RF), and extreme gradient boosting (XGBoost) were tested on the Dryad dataset. LR models estimate the relationship between binary dependent variables and independent variables [28]. RF models utilize multiple trees, where each tree is constructed using a random subset of the data, and the outputs are aggregated to generate the final outcome [29]. XGBoost models utilize gradient boosting decision trees, where each tree sequentially corrects the error of its predecessor. It uses parallel and distributed computing, which makes it faster and scalable [30]. LR was used as a baseline model due to its ability to model binary predictions, whereas RF and XGB are tree-based models that offer robust performance when paired with tabular data. They can also capture non-linear relationships that may exist in the data. XGBoost was observed to yield the best performance upon testing and was finalized for NAFLD prediction. Deep learning methods were not tested since they require much larger datasets and are harder to interpret, which limits their use in clinical settings.

#### 2.5.2 Stage 2: Comorbidity Prediction

Three algorithms—LR, RD, and XGBoost — were also tested for comorbidities using a comorbidity-labeled NAFLD-positive dataset. A shared approach using a MultiOutputClassifier, where a single model predicts all three comorbidities, was initially attempted; however, for interpretability reasons, it was replaced with three separate models, one for each comorbidity. Switching to a three-model approach also allowed model-specific hyperparameter tuning if needed. Three XGBoost models were finalized, and polynomial features (using the PolynomialFeatures function with a degree of 2) were introduced to capture pairwise feature interactions.

Models were evaluated using commonly used evaluation metrics, including accuracy, precision, recall (also known as sensitivity), F1 Score, and ROC-AUC (area under the receiver operating characteristic curve). Evaluation was done on an 80-20 train-test split.

### 2.6 Model Explainability

To address the need for transparency, which is necessary in a clinical tool, SHAP was used to interpret NAFLD model predictions. SHAP assigns a feature contribution score to each feature per prediction and displays them as bar plots with feature contribution reducing from top to bottom.

Since comorbidities are primarily dependent on threshold features for their prediction, a radar plot was used to enhance their explainability. Each axis of the plot represents a key parameter (threshold feature), and the radar extends further towards specific features if they have elevated values. Risk assessment was also calculated based on the probability of predictions, and color-coding was applied to easily distinguish the risk levels.

Green indicates low risk (<30%), yellow indicates moderate risk (30–70%) and red indicates high risk (>70%).

### 2.7 Tool Deployment

The complete ML pipeline was deployed as a web-based tool by the Streamlit framework. The interface accepts 14 input fields - Gender, Age, BMI, WC, ALT, AST, GGT, HDL, TC, TG, HbA1c, FPG, SBP, DBP, calculates derived features based on the input values (after applying necessary unit conversion), and outputs NAFLD binary prediction along with probability of prediction and SHAP plot. If the NAFLD value is positive, the model tool also outputs binary predictions of each comorbidity with risk assessment (for quick understanding), probability of prediction, and a radar plot. All models are saved using joblib, a Python library used for lightweight pipelining, and loaded at runtime to avoid retraining during different user interactions. The entire pipeline has been designed with a modular framework enabling flexibility to train models with different, larger datasets if available.

## 3. RESULTS

### 3.1 NAFLD Prediction Model Performance

The three models were initially trained with the original dataset to see if synthetic data generation could be avoided. As shown in Table 2, model performances were suboptimal, especially in recall, due to the limited number of NAFLD-positive cases.

**TABLE 2:** Model performance without synthetic data generation.

| **Model** | **Accuracy** | **Precision** | **Recall** | **F1 Score** | **ROC-AUC** |
| --- | --- | --- | --- | --- | --- |
| **LR** | 87.60 | 70.92 | 49.81 | 58.52 | 91.64 |
| **RF** | 87.90 | 72.09 | 50.76 | 59.57 | 90.61 |
| **XGB** | 87.56 | 68.70 | 53.63 | 60.24 | 90.19 |

Since the models were inefficient in correctly classifying the majority of NAFLD-positive cases without class balancing, synthetic data generation was necessary. A rule-based method was applied, and Table 3 shows the performance of models after retraining with the augmented dataset. Improvement in performance was observed, with XGBoost yielding comparatively better results; hence, it was selected as the final model for NAFLD prediction.

**TABLE 3:** Model performance with synthetic data generation.

| **Model** | **Accuracy** | **Precision** | **Recall** | **F1 Score** | **ROC-AUC** |
| --- | --- | --- | --- | --- | --- |
| **LR** | 90.79 | 92.74 | 87.18 | 90.19 | 97.01 |
| **RF** | 91.07 | 93.32 | 87.77 | 90.46 | 97.08 |
| **XGB** | 91.17 | 94.07 | 87.97 | 90.89 | 97.24 |

While the final model performs well, further improvements in recall (actual positives predicted as negatives) could be obtained through the use of higher-quality datasets, if available.

### 3.2 Comorbidity Prediction Models

Each comorbidity model was trained independently using XGBoost, and the tree-based models achieved a perfect fit (Tables 4,5, and 6), which is potentially due to the size of the dataset used for training.

**TABLE 4:**
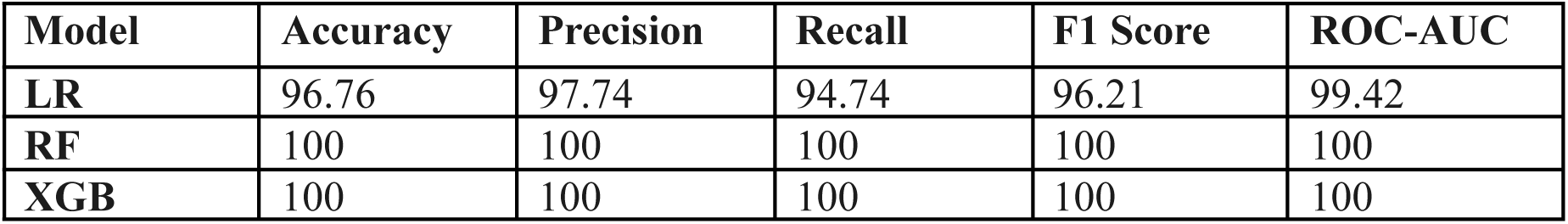
Performance of models for predicting hypertension.

**TABLE 5:** Performance of models for predicting CVD.

| **Model** | **Accuracy** | **Precision** | **Recall** | **F1 Score** | **ROC-AUC** |
| --- | --- | --- | --- | --- | --- |
| **LR** | 92.37 | 93.70 | 95.26 | 94.48 | 97.91 |
| **RF** | 100 | 100 | 100 | 100 | 100 |
| **XGB** | 100 | 100 | 100 | 100 | 100 |

**TABLE 6:**
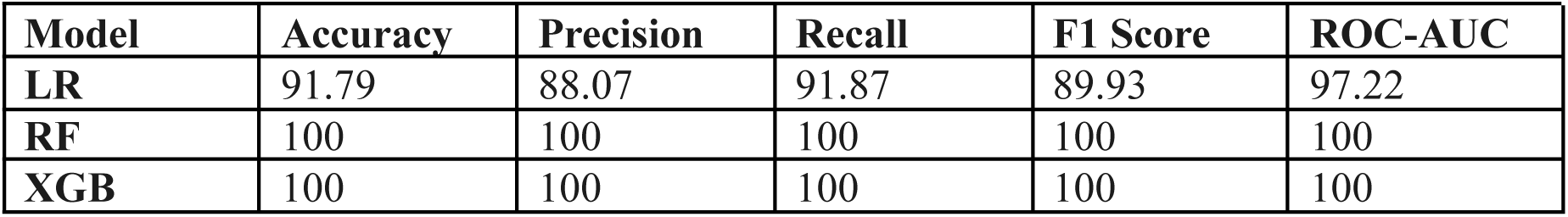
Performance of models for predicting Prediabetes.

| **Model** | **Accuracy** | **Precision** | **Recall** | **F1 Score** | **ROC-AUC** |
| --- | --- | --- | --- | --- | --- |
| **LR** | 91.79 | 88.07 | 91.87 | 89.93 | 97.22 |
| **RF** | 100 | 100 | 100 | 100 | 100 |
| **XGB** | 100 | 100 | 100 | 100 | 100 |

Despite the perfect metrics, the XGB model was selected for comorbidities, allowing for the potential use of a larger, more diverse dataset in the future, if available. The model was tested out using different values that represent multiple scenarios of positive and negative comorbidity cases, and the performance was observed to be robust.

### 3.3 Explainability and Visualization

SHAP plots were generated for individual predictions to enhance model transparency (Figure 2), along with NAFLD status and the corresponding prediction probability. They rank the top contributing features, and METS-IR, ALT, TG, TyG, and BMI were consistently identified as the top drivers. The bar plots enable clinicians to easily identify which feature drives the model prediction for each patient.

**Figure 2.**
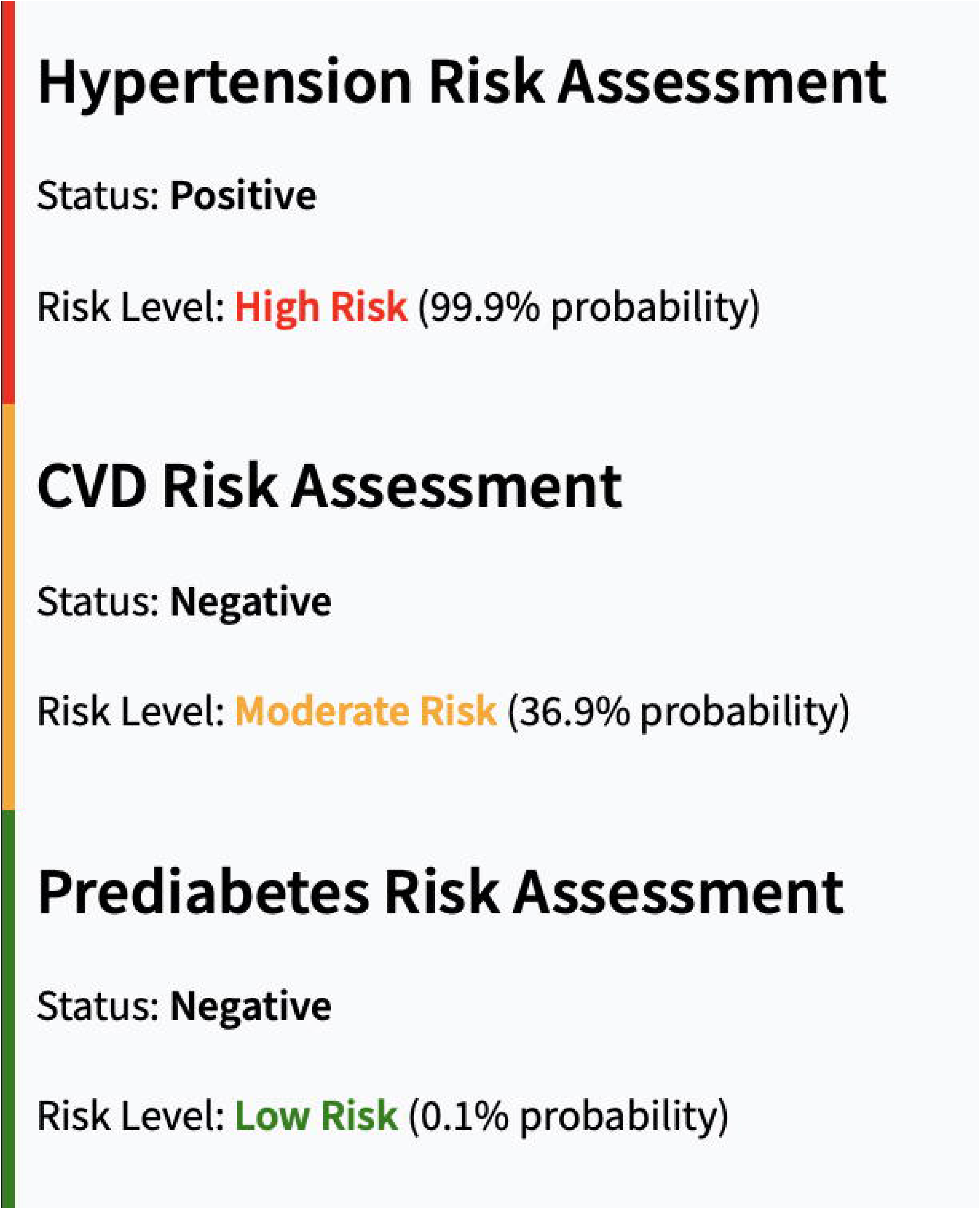
SHAP summary bar plot of top clinical features contributing to NAFLD prediction.

For patients listed as having NAFLD, the tool screens for comorbidities and outputs a color-coded risk assessment of each comorbidity, along with the predicted probability (Figure 3) and radar plots visualizing deviations of key biomarkers from normal thresholds (Figure 4). Simplifying the visualization of all three comorbidities into a single figure allows better understanding and keeps the interface clutter-free.

**Figure 3.**
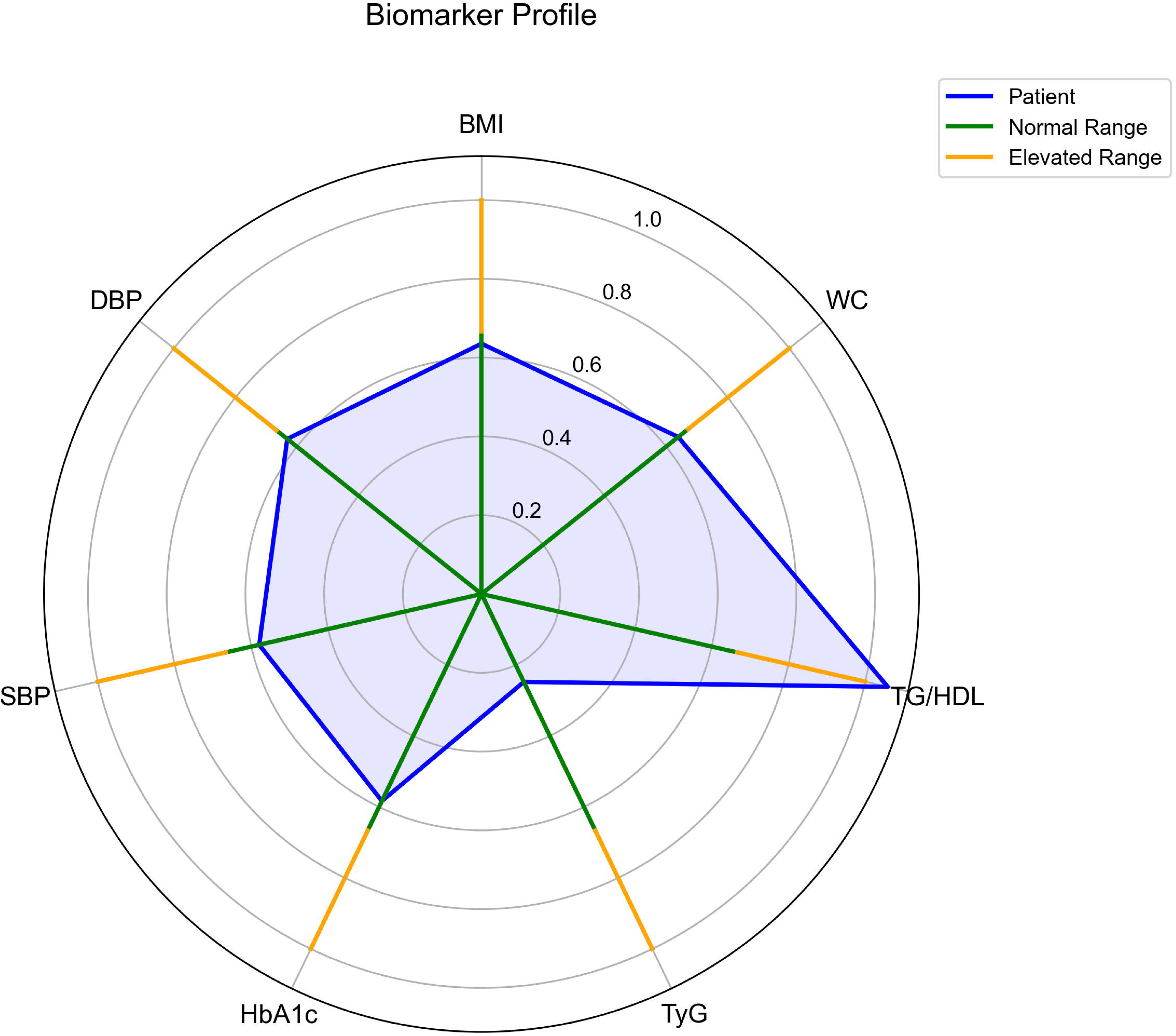
Color-coded risk assessment of comorbidities with associated probabilities.

**Figure 4.**
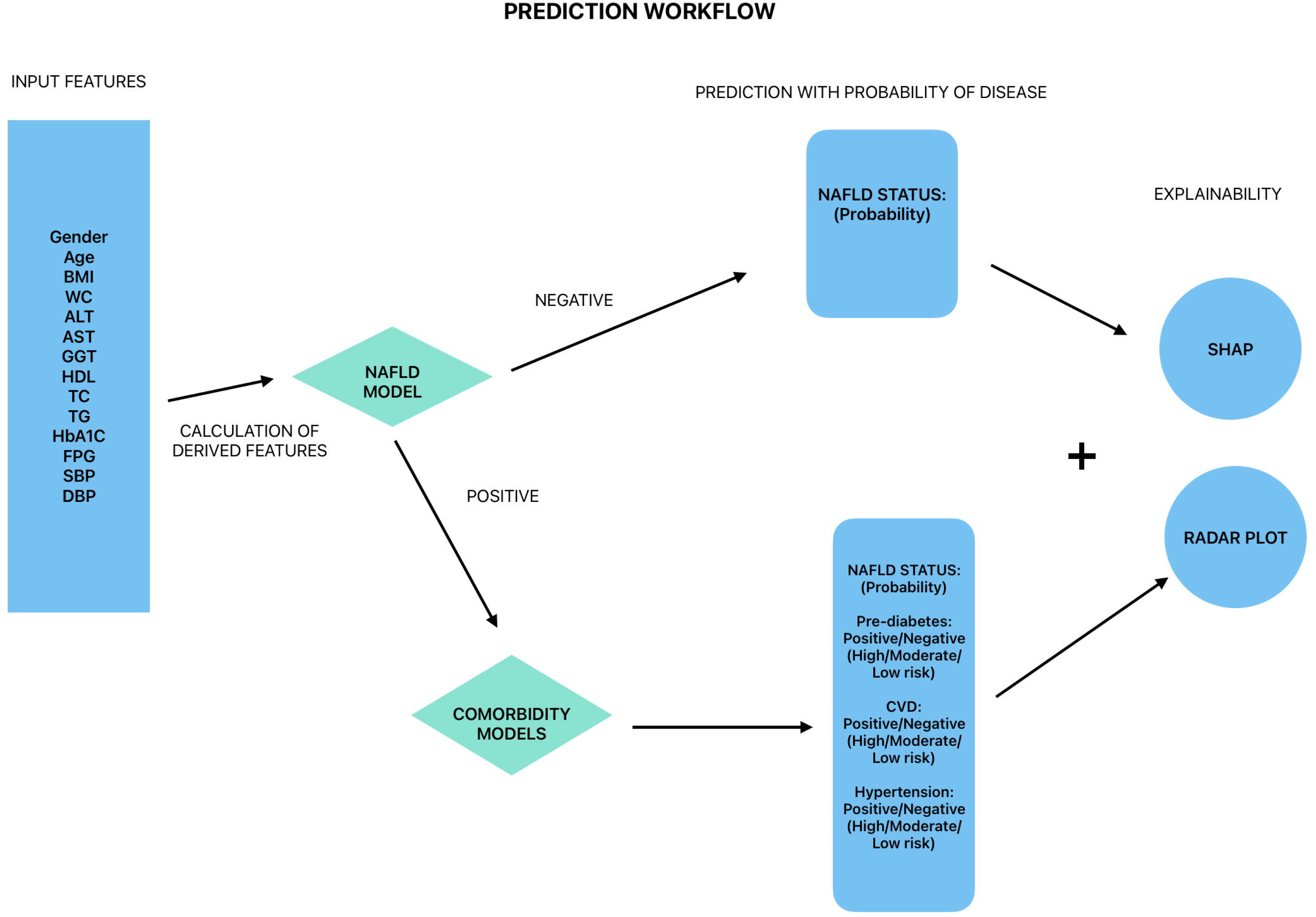
Radar plot showing elevated TG\HDL range compared to normal diagnostic thresholds.

### 3.4 Tool Integration

The Streamlit interface integrates the two-stage pipeline into a single, user-friendly tool, allowing clinicians to enter inputs and view the corresponding outputs. (Figure 5) shows the complete workflow of the tool from input data to visualization.

**Figure 5.**
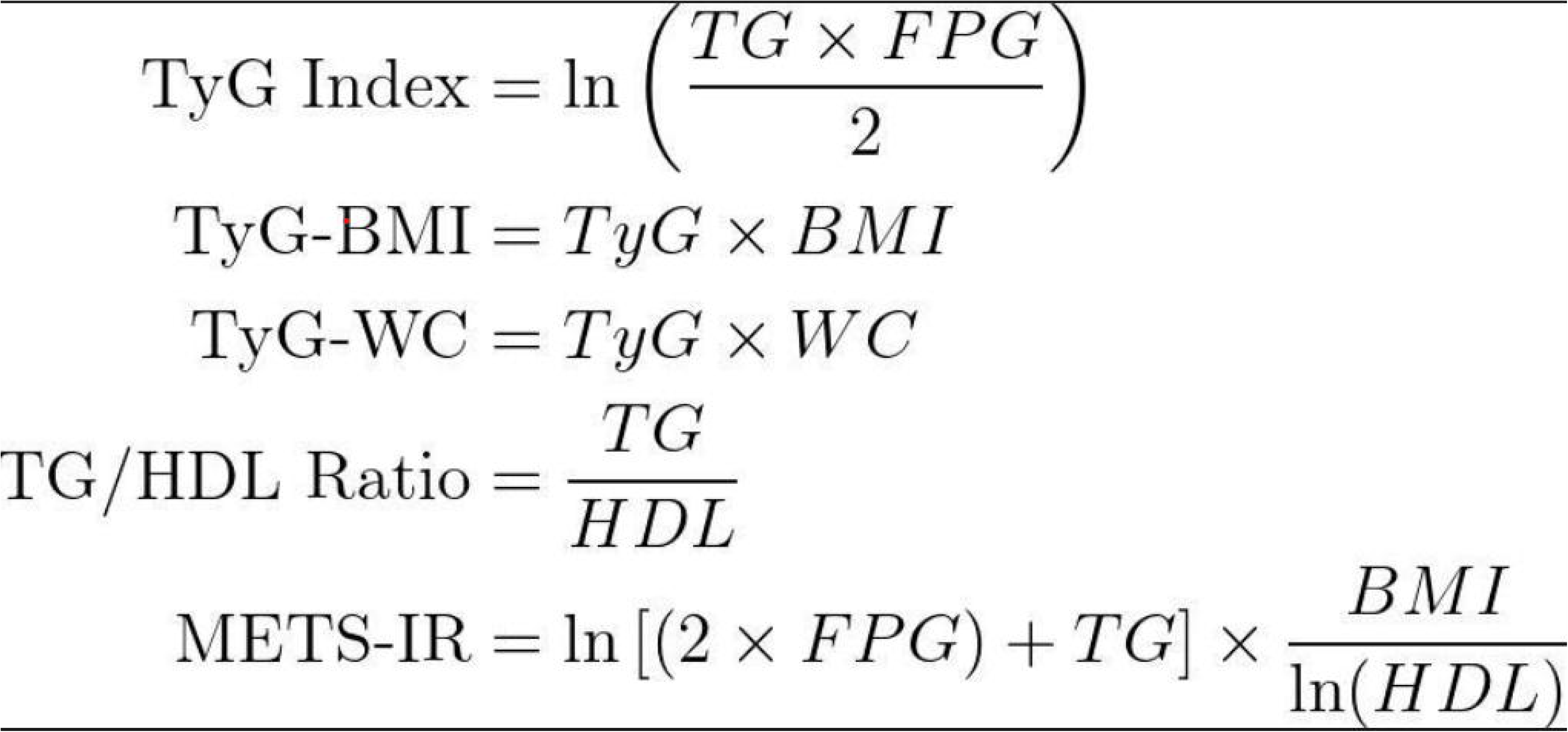
Workflow diagram of the prediction tool illustrating processes from feature input to NAFLD detection, comorbidity evaluation, and outputs including SHAP explanations and radar plots

## 4. DISCUSSION

NAFLD is a globally prevalent health condition that affects about one-third of the population while often being asymptomatic. This study demonstrates the development of an interpretable, ML-based diagnostic pipeline for NAFLD and related comorbidities using routinely collected, non-invasive clinical data, intended for use as a diagnostic aid in primary care. The pipeline is built around a two-stage framework. It integrates XGBoost models with SHAP and radar plots for visualization, making it a modular, accessible, inexpensive, and easy-to-operate solution.

The results from NAFLD models, after synthetic data augmentation, with an accuracy of 91.17%, precision of 93.32%, F1 Score of 90.19%, and ROC-AUC of 97.01%, are robust. The synthetic data generation is limited to clinically observed ranges, which helps keep it realistic. The use of derived metabolic indices such as the TyG index, TyG-BMI, and METS-IR further enhances the predictive ability of the model. While the recall of 87.77% is strong, there is potential to further improve it with higher-quality, better-balanced datasets on availability. Notably, the model leverages non-invasive features captured during routine clinical visits, making it suitable for use in resource-limited environments and primary care facilities without specialized imaging equipment. This strengthens the aim of the tool to serve as a screening aid for clinicians, enabling them to flag high-risk patients for further diagnosis or treatment.

In addition to detecting NAFLD, the tool also focuses on identifying three prevalent comorbidities associated with NAFLD, including CVD, Prediabetes, and Hypertension. Since NAFLD shares a bidirectional relationship with these diseases, this allows for the assessment of both the disease and its comorbidities as a holistic system. This is important because comorbidities contribute to the faster progression of NAFLD into severe stages. Both tree-based comorbidity models (RF and XGBoost) achieved perfect performance (100% accuracy, precision, recall, F1 score, and AUC-ROC). In contrast, the LR model shows an accuracy of 96.76, a recall of 84.74, and an ROC-AUC of 99.42 for Hypertension, an accuracy of 91.79, a recall of 91.87, and an ROC-AUC of 97.22 for Prediabetes, and an accuracy of 91.79, a recall of 95.26, and an ROC-AUC of 97.91 for CVD. XGBoost was selected as the prediction model because different values relating to comorbidities were manually tested to evaluate each model, and XGBoost demonstrated robust performance. However, the perfect metrics might be due to the limited size of the dataset, with each comorbidity having fewer than ideal representation, allowing advanced models like XGBoost to learn it perfectly. This is acknowledged as a limitation of the dataset. The pipeline was designed to be modular, allowing for the possibility of replacing the comorbidity dataset with larger, more diverse data in the future. Synthetic data generation was not considered, as the labels for each comorbidity were limited in number, and synthetic entries risked deviating from real-world cases. The XGBoost model was still preferred, considering that studies have shown tree-based models to perform better in capturing diverse, non-linear relationships in tabular data [31]. Three separate XGBoost models were used, one for each comorbidity, to allow potential fine-tuning specific to each model if required.

The explainability aspect strengthens the tool as a primary care aid. The integration of SHAP allows clinicians to trace model outputs to specific biomarkers. Radar plots, added to comorbidity predictions, bring together all key biomarkers (selected based on those used by clinicians to predict either of these comorbidities) in a single plot, helping users readily understand which feature led to the predictions. Current cutoffs are based on clinically adopted thresholds; however, future upgrades may include patient-specific baselines.

Streamlit’s deployed user interface is easy to use; the user simply enters input values in the specified units on the interface and clicks the "Predict" button to obtain output results along with their visualizations. The models are saved using the joblib library, eliminating the need to retrain the model every time the user runs the tool.

The lack of data for external validation remains a limitation of the study. Before deployment in healthcare fields, the tool must be validated using a different, larger, and more diverse dataset. Synthetic data generation can also be avoided if a well-balanced dataset of sufficient size is available. Future studies should also consider longitudinal data that allows for tracking changes in patients over specific periods for stagewise detection and include additional factors, such as diet, exercise, and medication use, which may enhance the diagnosis.

## 5. CONCLUSION

This study presents an AI-assisted diagnostic tool designed to predict Non-Alcoholic Fatty Liver Disease (NAFLD) and related comorbidities, including Cardiovascular Disease (CVD), Hypertension, and Prediabetes, using non-invasive, comparatively inexpensive, and routine clinical data. It utilizes an XGBoost model for prediction and integrates explainability through SHAP and radar plots to produce a practical tool that can aid clinicians in primary care settings, especially in areas without specialized diagnostic equipment.

The two-stage predictive pipeline focuses on NAFLD detection in the first stage and offers robust predictive performance. Synthetic data was generated for class balancing in the Dryad dataset used for NAFLD predictions. The positive NAFLD cases are diagnosed for the presence of comorbidities, with the models trained on a separate dataset, which is a subset of the Dryad dataset containing only NAFLD-positive cases. However, the comorbidity models, while yielding robust results on tests, exhibit perfect values on evaluation metrics, possibly due to the limited dataset size and threshold-based labeling. A larger, more diverse dataset is needed for further testing on this part.

The explainability aspect, which is vital for adopting the tool into primary healthcare, is obtained through SHAP-based feature attributions and radar plots that show elevations in threshold features. This addresses the ‘whys’ behind the prediction and helps reduce the black box nature of the model. The pipeline, along with visualizations, was integrated into an easy-to-operate tool using Streamlit for deployment, which allows accessibility within a limited computational setup, making it ideal for rural, low-resource settings.

The study acknowledges limitations including dataset size, external validation, and lack of longitudinal data tracking for a more dynamic progress detection. Future work should focus on using larger, more diverse datasets for comorbidities, including longitudinal data, and explore the integration of additional comorbidities associated with NAFLD, such as polycystic ovary syndrome (PCOS), sleep apnea, and chronic kidney disease (CKD). Refining the user interface for integration into electronic health records (EHR) to screen large amounts of patient data in ways that allow continual learning to keep the model continually improving is also a future direction.

In short, this work demonstrates the potential of data-driven, explainable AI integrated systems to assist with early detection and management of NAFLD and its related comorbidities to offer a cost-effective, easy-to-operate, and transparent approach that can complement the existing clinical practices.

## Data Availability

All data produced in the present study are available upon reasonable request to the authors.

## DATA AVAILABILITY

The datasets generated and/or analyzed and the code used for analysis during the current study are available from the author on reasonable request.

